# Impact of the home confinement related to COVID-19 on the device-assessed physical activity and sedentary patterns of Spanish older adults

**DOI:** 10.1101/2020.11.19.20234583

**Authors:** Ángel I. Fernández-García, Jorge Marin-Puyalto, Alba Gómez-Cabello, Ángel Matute-Llorente, Jorge Subías-Perié, Jorge Pérez-Gómez, Gabriel Lozano-Berges, Asier Mañas, Amelia Guadalupe-Grau, Marcela González-Gross, Ignacio Ara, José A. Casajús, Germán Vicente-Rodríguez

**Affiliations:** GENUD (Growth, Exercise, Nutrition and Development) research group, University of Zaragoza, Zaragoza, Spain; Faculty of Health and Sport Science (FCSD, Ronda Misericordia, 5, 22001-Huesca, Spain). Department of Physiatry and Nursing, University of Zaragoza, Spain; Red española de Investigación en Ejercicio Físico y Salud en Poblaciones Especiales (EXERNET); Instituto Agroalimentario de Aragón -IA2- (CITA-Universidad de Zaragoza); Centro Universitario de la Defensa, Zaragoza, Zaragoza, Spain; Centro de Investigación Biomédica en Red de Fisiopatología de la Obesidad y Nutrición (CIBERObn); Faculty of Health, Department of Physiatry and Nursing, University of Zaragoza, Zaragoza, Spain; HEME (Health, Economy, Motricity and Education) research group. University of Extremadura, Cáceres 10003. Spain; GENUD Toledo Research Group, Universidad de Castilla-La Mancha, Toledo, Spain; CIBER of Frailty and Healthy Aging (CIBERFES), Madrid, Spain; ImFine Research Group. Facultad de Ciencias de la Actividad Física y del Deporte-INEF, Universidad Politécnica de Madrid, Madrid, Spain

**Keywords:** Aging, coronavirus, elderly, exercise, home-quarantine, inactivity, pandemic

## Abstract

**Objectives:** The main objective of this study was to device-assess the levels of physical activity and sedentary behaviour patterns of older adults during the situation prior to COVID-19 pandemic, home-quarantine and the ending of isolation. We also aimed analysing the effectiveness of an unsupervised home-based exercise routine to counteract the potential increase in sedentary behaviour during the periods within the pandemic.

**Methods:** 18 non-institutionalized elderly (78.4±6.0 y.), members of the Spanish cohort of EXERNET-Elder 3.0 project participated in the study. They were recommended to perform an exercise prescription based on resistance, balance and aerobic exercises during the pandemic. Wrist triaxial accelerometers (ActiGraph GT9X) were used to assess the percentage of sedentary time, physical activity and sedentary bouts and breaks of sedentary time. An ANOVA for repeated measures was performed to analyse the differences between the three different periods.

**Results:** During home-quarantine, older adults spent more time in sedentary behaviours (71.6±5.3%) in comparison with either the situation prior to the pandemic (65.5±6.7%) or the ending of isolation (67.7±7.1%) (all *p*<0.05). Moreover, participants performed less bouts of physical activity and with a shorter duration during home quarantine (both *p*<0.05). Additionally, no differences in the physical activity behaviours were found between the situation prior to the pandemic and the ending of isolation.

**Conclusions:** According with our results, the home-quarantine could negatively affect health due to increased sedentary lifestyle and the reduction of physical activity. Therefore, our unsupervised exercise program does not seem to be a completely effective strategy at least in this period.

**What is already known on this topic:**

- Although the available information is scarce and includes subjective methodology (questionnaires), it seems that the COVID-19 pandemic has negatively affected physical activity patterns.
- It is known that physical activity interventions are effective in improving health and reducing sedentary lifestyle in older adults. Nevertheless, little is known about whether an unsupervised home-based exercise routine is an effective alternative to counteract the potential increase in sedentary behaviour in this specific population during the pandemic lockdown.

**What are the findings? / What this study adds:**

- Despite unsupervised training, during home-quarantine, older adults spent more sedentary time than in the situation prior to COVID-19 and the ending of isolation (phase 0).
- There were no differences in break of sedentary time patterns between the situation prior to COVID-19 and the periods within the pandemic.
- During home-quarantine older adults performed fewer and shorter physical activity bouts than in the situation prior to COVID-19 despite unsupervised training.
- Our unsupervised home-exercise routine was not a completely effective alternative to avoid the increase of sedentary behaviour during home-quarantine.

**How might it impact on clinical practice in the future?:**

- Our findings can be used as a starting point to manage isolation restrictions more effectively and to develop strategies to promote physical activity and reduce sedentary behaviour among older adults during situations of forced lockdowns, as in the present COVID-19 pandemic.

## 1. Introduction

The coronavirus disease-19 (COVID-19) pandemic is an unprecedented health crisis that has forced millions of people to live in home-confinement. Quarantine was one of the main government actions to reduce the risk of spread. In Spain, the lockdown lasted 98 days, including the different stages of de-escalation with specific restrictions until the so-called “new normality”. Unavoidably, these restrictions are repeating in the present COVID-19 world situation and have modified the routine activities by increasing sedentary time[1,2], so the pandemic may carry considerable risks to health and well-being[3–8]. Furthermore, COVID-19 spread is especially important in people at increased risk for severe illness like older adults.

Sedentary behaviour (SB), defined as any waking behaviour characterized by an energy expenditure <1.5 METs[9], has been considered by some research as a new risk factor among the older adults, even regardless of physical activity (PA)[10]. SB is related to an increased risk of cardiovascular disease, diabetes, mental health problems and some types of cancer, as well as premature all-cause, cancer and cardiovascular disease mortality[11–14]. On the other hand, an increase in PA levels has been proposed as a relevant strategy to achieve successful ageing[15–17] due to its positive health and fitness benefits[18,19]. Nevertheless, older adults are characterized by a very sedentary lifestyle (only over 20% follow the PA recommendations)[20]. The most widely used tool to measure PA and SB are the self-reporting questionnaires[21,22], although it is known that older adults tend to underestimate the time spent in sedentary activity and overestimate PA levels when subjective measurements are compared with accelerometers[23,24].

Given the negative health consequences of reducing PA and increasing SB, from the beginning of the COVID-19 outbreak some authors proposed continuing PA at home to stay healthy in the current precarious environment[7,25]. Such forms of exercise may include strengthening exercises, balance activities, walking at home, stretching, or a combination of all or some of them[7]. Considering all the above, a home-based exercise routine prescription based on safe, simple and easily implementable exercises in reduced spaces, could be an effective strategy to prevent or attenuate the effects of the potential increase in inactivity following the enforced lockdown in this population.

The impact of home-confinement caused by the COVID-19 pandemic on PA and SB of older adults has yet to be investigated in depth and the few studies that exist have been carried out with subjective methods. Therefore, the main aim of this study was to evaluate the differences in PA, SB and break of sedentary time (BST) between the situation prior to COVID-19 pandemic, home-quarantine (HQ) and the ending of isolation (EI): phase 0, analysing the effectiveness of an unsupervised home-based exercise routine to combat the potential increase in SB during the pandemic.

## 2. Methods

### 2.1 Participants

The inclusion criteria were being older than 65 years and not suffering from cancer or dementia. A total of 24 non-institutionalized older adults (14 women; 78.4±6.0 y.), members of a subgroup of the Spanish cohort of EXERNET-Elder 3.0 project, accepted the invitation to participate in the study. All of them were performing since January 2020 until the lockdown, a supervised multicomponent exercise program three days per week.

Participants were not involved in the design, or conduct, or reporting, or dissemination plans of our research

### 2.2 Physical Activity and Sedentary Behaviour assessment

All participants were familiar with accelerometry, anyway they were instructed by phone and in written to continuously wear the accelerometer on their non-dominant wrist. Wrist accelerometers have shown to increase adherence to wear protocols [26]. They also wore them when sleeping and removing it only for water-based activities in three different periods: 1) Usual lifestyle (UL) prior to COVID-19 pandemic (December 2019). During this period, the participants were asked to maintain their daily routines unchanged for seven consecutive days; 2) HQ: the last two days of home-quarantine (14-15 May 2020). In this period, participants stayed at homes, and they were only allowed to leave them for medical appointments or shopping for basic needs; 3) EI: First two days of “phase 0” of the ending of isolation (16-17 May 2020). At this point the quarantine was lifted for three hours per day, although older adults could go out only one hour.

PA and SB were evaluated with an ActiGraph GT9X triaxial accelerometer (ActiGraph GT9X Link; Actigraph, 49 E. Chase St. Pensacola, FL 32502) and the data were analysed with the ActiLife software (ActiLife v6.13.3, ActiGraph Corp., Pensacola, Florida). Accelerometer data were collected at 60-Hz and were later aggregated into 60-second epochs. Non-wear time was defined by and interval of at least 60 consecutive minutes of zero activity intensity counts, with an allowance for 1-2 minutes of counts between 0 and 100[27]. Sleeping periods, non-wear time, and any day with less than 10 hours of wear time were removed from the analysis. To be included in the sample, participants were required to have at least one valid day in all evaluation periods[28]. A SB was considered when the monitor registered <1853 counts per minute[28], while data ≥1853 counts was included as PA. In a further analysis of SB, three different aspects were evaluated: the percentage of wear time spent in SB, bouts of SB which were defined as periods of at least 10 consecutive minutes of SB and BST, which were considered as any interruption of SB independently of its duration. Regarding bouts of PA, a minimum block of 30 consecutive minutes was required, as the minimum estimated duration of the home-based exercise session.

### 2.3 Implementation and adherence of unsupervised exercise training

Upon delivery of the accelerometers during HQ by express postal delivery service, the participants received on paper a voluntary home-based exercise routine prescription, which was recommended to be performed five days per week, from last week of home-confinement until the end of de-escalation process. It consisted on seven resistance exercises, four for lower (chair squat, hip abduction and adduction and calf raise) and three for upper limbs (push-ups on the wall, biceps curl and shoulder abduction) carried out in two sets of 12-15 repetitions using the resistance of body weight and lifting home-available light free weights as liquid bottles; three balance exercises (semi-tandem or tandem position, single foot stand and dynamic balance heel-toe) executed twice along 20 seconds, two times each one; and 20-30 minutes walking performed in one or several sets of 10 minutes minimum. The resting time between sets and exercises was 45 seconds.

Furthermore, participants registered if they performed the training session during the days in which they had the accelerometers.

### 2.4 Statistical analysis

Normality of the sampling distribution was proved using Shapiro-Wilk test and those variables which did not follow a normal distribution were transformed using the square root and the reverse function. Statistical significance was set at level *p*<0.05 in all tests. Descriptive statistics (mean ± standard deviation (SD)) were calculated for all variables and an ANOVA for repeated measures was done to examine the differences in SB, PA and sedentary breaks between periods. For those variables transformed to normalized, the original mean and SD values were reported. Finally, a t-test for related samples was done to compare the percentage of time spent in PA between the schedule in which older adults could leave their homes, and the rest of time when they stayed at home during EI period.

All the analyses were performed using the Statistical Package for Social Sciences software (SPSS, v. 25.0 for WINDOWS; SPSS Inc., Chicago, IL, USA), and values of *p*<0.05 were considered statistically significant.

The protocol of the EXERNET-Elder 3.0 study has been approved by the Ethics Committee of Clinical Research from the Alcorcón Foundation University Hospital (16/50), additionally the Research Ethics Committee of the Autonomous Community of Aragon approved the specific study during the home-quarantine (nº 10/2020). The whole study followed the ethical guidelines of the Declaration of Helsinki 1964, revised in Fortaleza (2013). Participants received written information detailing the purpose, procedures, benefits and risks that might result from their participation. All who voluntarily agreed to participate signed an informed consent.

## 3. Results

### 3.1 Device-assessed data

Data from six participants could not be included in the analysis due to delay in delivery of accelerometers by the postal delivery service, so finally data from 18 older adults (11 women) were included. Participants wore the accelerometers for an average of 6.9±0.2 days (1046.2±86.1 minutes per day) in UL, 1.7±0.5 days (936.1±146.8 minutes per day) in HQ and 2.0±0.0 days (1066.4±130.0 minutes per day) in EI. Table 1 shows the results of the analysed variables.

**Table 1.** Participants’ SB and PA patterns in the different evaluation periods.

|  | Usual lifestyle<br>(UL) | Home-quarantine<br>(HQ) | Ending of isolation:<br>phase 0 (EI) |
| --- | --- | --- | --- |
| **SB (% total wear time)** | 65.5 ± 6.7 | 71.6 ± 5.3<sup>ac</sup> | 67.7 ± 7.1 |
| **BST (number/h)** | 5.0 ± 0.5 | 4.6 ± 0.8 | 4.6 ± 0.8 |
| **10-min SB bouts (number/h)** | 0.9 ± 0.1<sup>bc</sup> | 1.1 ± 0.2 | 1.1 ± 0.2 |
| **AVG time of 10-min SB bouts**<br>**(min/bout)** | 30.4 ± 5.5 | 32.0 ± 8.9 | 29.4 ± 7.3 |
| **30-min PA bouts (number/day\*)** | 3.2 ± 1.7 | 2.2 ± 1.9<sup>ac</sup> | 3.2 ± 2.1 |
| **AVG time of 30-min PA bouts**<br>**(min/bout)** | 41.4 ± 5.2 | 31.7 ± 15.6<sup>a</sup> | 42.1 ± 9.09 |

*Note:* SB: sedentary behaviour; BST: break sedentary time; AVG: average; PA: physical activity; \*adjusted by 24 valid hours; <sup>a</sup> Significant difference with UL; <sup>b</sup> Significant difference with HQ; <sup>c</sup> Significant difference with EI

During UL participants had fewer sedentary bouts than in HQ and EI (*p*<0.05), although there was no difference on its average duration among periods. Furthermore, in HQ older adults spent a higher percentage of wear time in SB compared with UL and EI (*p*<0.05), although no differences were found in BST. Regarding PA, during HQ elders performed fewer bouts and their average duration was shorter than in UL (*p*<0.05).

### 3.2 Adherence of unsupervised home-based exercise program

Regarding participants’ compliance of the unsupervised home-based exercise routine during HQ, 22.2% of older adults carried it out both days, 39.9% only one and the other 38.9% did not perform it. In EI period only 11.1% performed the training prescription two days, while 50% only one and the other 38.9% did not carry out any session.

### 3.3 Physical Activity during periods were restrictions were lifted in Ending Isolation

Additionally, during the two days of EI, 44.4% of older adults went out of home both days during the schedule in which restrictions were lifted, while 27.8% went out only one day and the other 27.8% did not go out any day. Furthermore, the average time spent in PA during the periods in which older adults were able to leave their homes, were significantly higher than when restrictions were activated during EI (49.2±14.2% vs. 29.0±7.5%; *p*<0.001).

## 4. Discussion

The main findings of this study are: (i) Despite unsupervised training, during home-quarantine, elders spent more sedentary time than in usual lifestyle and during the ending of isolation; (ii) there were no differences in the breaks of sedentary time patterns between usual lifestyle and the periods within the pandemic; (iii) during home-quarantine older adults performed fewer and shorter PA bouts than in the usual lifestyle despite unsupervised training.

### 4.1 Changes in habits: Sedentary behaviour, Physical Activity and Breaks of Sedentary Time

To the best of our knowledge, no previous study has analysed through device-assessment the SB and PA patterns during the pandemic. The time spent in SB by our participants ranged between 66% of waking time in UL and 72% in HQ. Long periods of SB during UL ranging between 65-80% of waking time have also been reported in this population by a previous systematic review that analysed data obtained by accelerometry[29]. It is known that prolonged SB increases the risk of chronic diseases and mortality[10], so the increase of SB during HQ, could adversely affect health[10].

A recent study which aimed to analyse changes in activity and daily routine among Chinese citizens during home-quarantine, the prevalence of insufficient PA in this population rose over 4-fold during the epidemic quarantine (57.5%), compared with the non-epidemic period (14.1%) [4]. Moreover, the authors concluded that participants’ screen time increased as PA level declined in the HQ. Some of these conclusions could be extended internationally, since another recent research that obtained its data through an online international survey (of people aged over 18, including group over 55), concluded that the COVID-19 home-confinement had a negative effect on all PA intensity levels and additionally, increased daily sitting time in a 29% [2]. In our study, the increase of SB during the pandemic was much lower (6.1% in HQ; 2.2% in EI) in comparison with UL, although it must be considered that the population was different and previous study does not include specific data neither in adults or older adults. Also, low PA levels are associated with increased prevalence of anxiety [30], which reinforces the importance of the promotion of home-based PA during such lockdowns, to reach at least the recommended 150 min of PA per week[25].

Regarding BST, a similar result (5.3 breaks/hour) was obtained in a previous study with frail older adults in UL[31] and Dos Santos et al.[32] obtained a greater amount (174 breaks/day) in non-frail older adults, although the accelerometers were located on the hip in both studies. Previous research have shown that greater number of breaks in sitting behaviours is associated with better physical function[33] and longer SB bouts are detrimental to overall health[34]. Nevertheless, a recent study concluded that breaking-up sedentary time more often reduce frailty only in those older adults who are inactive[35]. Considering that older adults spent a longer time in SB during HQ in our study, the absence of differences between periods in BST (n/hour), could suggest that along SB in HQ, participants performed fewer BST than in the other two periods.

### 4.2 Adherence and effectiveness of unsupervised home-based exercise program

With regard to unsupervised training program, although it was not enough to avoid the differences with UL, it cannot be affirmed that has not helped to partially avoid a greater sedentary time or mitigate a possible decline of fitness levels, having a positive effect on health. Moreover, two aspects should be taken into consideration. Firstly, the difficulty of walking at home, especially for older people, and secondly, most of strength and balance exercises, had not increased the PA too much. The low compliance of the exercise program could be explained because unsupervised training was not motivating enough to create the necessary adherence in older adults, even though they were performing a supervised exercise program previously. This undoubtedly affected the results and could be the reason why the elderly did not maintain the SB patterns of UL during HQ. On the other hand, the decrease of SB and the greater number and longer duration of PA bouts in EI compared to HQ could be associated with the possibility of going out of home for an hour per day, rather than the fact of doing more sessions, given the low compliance of the program and the increase of PA time during the schedule in which restrictions were lifted. In this regard, the absence of differences between EI and UL in all variables except in the number of sedentary bouts per hour, should be taken with caution, because the data of EI was taken in the first two days when the restrictions were a partially lifted. Consequently, older adults could increase their PA due to their desire to go out for a walk, although this behaviour might not be maintained over time.

### 4.3 Future perspectives

Taking into account the adverse effects of sedentary lifestyle and the negative consequences of COVID-19 in PA and SB patterns of older adults, our findings can be used as a starting point to develop strategies to promote PA among older adults during similar situations of lockdowns. Further research is needed to analyse the effectiveness of implementing motivational strategies or semi-supervision in terms of improving adherence. Regarding fitness impact, although little is known about the dose-response relationship, previous systematic review has shown the greater benefits of supervised with respect to unsupervised strength and/or balance programs, being particularly prominent when compared with completely unsupervised[36]. The use of eHealth and exercise videos, which focuses on encouraging and delivering PA interventions through the Internet, mobile technologies, and television[37] are other viable avenues for maintaining physical function and mental health during this critical period[7]. A recent study has demonstrated, through Google Relative Search Rate, that the community interest in exercise surged following the lockdown[6]. Nevertheless, older adults tend to lag behind in their adoption of technology, so a good approach would be to get older adults to be able to use new technologies efficiently.

### 4.4 Strengths and limitations of this study

The current research is highly topical and relevant, since we have not yet defeated the pandemic. Quarantines and confinements are repeated in Spain and different parts of the world. To our knowledge, this is the first study that device-assessed the impact of home-confinement caused by COVID-19 in PA and SB patterns of older adults, while undergoing an unsupervised exercise intervention.

Nevertheless, some limitations of our study should be mentioned. Firstly, the small sample size limits the generalisation of the results and our findings should be interpreted considering the absence of a control group for ethical reasons. Secondly, although the attendance was registered, the level of compliance of exercise prescription could not be recorded because it was an unsupervised training. And thirdly, participants wore the accelerometer fewer days during the periods evaluated within the pandemic, although it is probable that, due to the restrictions imposed, daily routines were similar and there would be no differences between weekdays and weekends, as previous studies have shown in this population in a usual life situation[39]. Moreover, specific cut points for light, moderate and vigorous PA have not yet been defined for this population and body location for ActiGraph accelerometers, so given the importance of a deeper understanding of the PA patterns in healthy aging, future research effort should be made in this direction.

## 5. Conclusions and policy implications

According with our results, the home-quarantine increased the sedentary lifestyle in addition to causing a reduction of PA, which may lead to negative health effects in this population. Additionally, it seems that our unsupervised home-based exercise routine is not a completely effective strategy to maintain usual sedentary and PA patterns during the lockdown.

Quarantines and confinements that are currently happening in the world, show that the COVID-19 pandemic has not been defeated yet. Additionally, health recommendations to limit exposure and stay at home for as long as possible may increase the probability of semi-self-confinement in older people. Therefore, the situation stresses the need for more research looking for a health promotion guideline showing effective strategies in terms of maintaining daily PA routines, especially in older adults, the most exposed group to excessive SB.

## Data Availability

Data are available upon reasonable request. Contact with the PI of the project G.V-R (ORCID identifier 0000-0002-4303-4097)

## Author contributions

Conceptualization, Á.I.F.-G., A.G.-C., J.A.C. and G.V.-R.; methodology, Á.I.F.-G., J.M-P, A.G.-C. and G.V.-R.; resources, Á.I.F.-G., J.M-P, A.G-C and A.M-L.; investigation: Á.I.F.-G, J.S-P. and G.V.-R.; data analysis: Á.I.F.-G., J.M-P and G.V.-R.; writing—original draft preparation, Á.I.F.-G., A.G.-C. and G.V.-R; writing—review and editing, Á.I.F.-G., J.M-P, A.M-L, J.A.C., A.G-G, and G.V.-R; visualization, A.M-L, J.S-P., M.G-G and J.A.C.; supervision, A.G.-C., J.A.C. and G.V.-R.; project administration, G.V.-R., J.A.C. and A.G.-C.; funding acquisition, G.V.-R., J.A.C. and A.G.-C. All authors have read and agreed to the published version of the manuscript.

## Acknowledgements

The authors are grateful to all collaborators: the nursing homes, health centers and participants involved in EXERNET-Elder 3.0 project and council social services, whose cooperation and dedication made this study possible. A. I. F. G has received a PhD grant from the Spanish Government (BES-2017-081402).

## Abbreviations

COVID-19: Coronavirus disease 19
MET: metabolic equivalent of a task
PA: physical activity
SB: sedentary behaviour
BST: break of sedentary time
UL: usual lifestyle
HQ: home-quarantine
EI: ending of isolation (phase 0)
SD: Standard deviation

